# Improving care for high impact users of hospital emergency departments: a mixed-method evaluation of a regional quality improvement programme ‘Supporting High impact users in the Emergency Department’ (SHarED)

**DOI:** 10.1101/2023.04.17.23287910

**Authors:** Carlos Sillero-Rejon, Megan Kirbyshire, Rebecca Thorpe, Gareth Myring, Clare Evans, Johanna Lloyd-Rees, Angela Bezer, Hugh McLeod

## Abstract

**Background:** The need to better manage frequent attenders or high-impact users (HIUs) in hospital emergency departments (EDs) is widely recognised. These patients often have complex medical needs and are also frequent users of other health and care services. The West of England Academic Health Science Network launched its Supporting High impAct useRs in Emergency Departments (SHarED) quality improvement programme to spread a local HIU intervention across six other EDs in five Trusts.

**Aim:** SHarED aimed to reduce ED attendance and hospital admissions by 20% for enrolled HIUs. To evaluate the implementation of SHarED, we sought to learn about the experience of staff with HIU roles and their ED colleagues, and assess the impact on HIU attendance and admissions.

**Methods:** We analysed a range of data including semi-structured interviews with 10 HIU staff; ED staff training; an ED staff experience survey; and ED attendances and hospital admissions for 148 HIUs enrolled in SHarED.

**Results:** Staff with HIU roles were unanimously positive about the benefits of SHarED for both staff and patients. SHarED contributed to supporting ED staff with patient-centred recommendations and provided the basis for more integrated case management across the health and care system. 55% of ED staff received training. There were improvements in staff views relating to confidence, support, training, and HIUs receiving more appropriate care. The mean monthly ED attendance per HIU reduced over time. Follow-up data for 86% (127/148) of cases showed a mean monthly ED attendances per HIU reduced by 33%, from 2.1 to 1.4, between the six months pre- and post-enrolment (p<0.001).

**Conclusion:** SHarED illustrates the considerable potential for a quality improvement programme to promote more integrated case management by specialist teams across the health and care system for particularly vulnerable individuals and improve working arrangements for hard-pressed staff.

**What is already known on this topic:** Frequent attendance in hospital emergency departments is a worldwide problem that, despite the national recognition of the rationale for better management of high-impact users, has relied on the local efforts of clinicians to change working practices.

**What this study adds:** The Supporting High impAct useRs in Emergency Departments (SHarED) quality improvement programme was successful in spreading a model of high-impact user management based on the identification, proactive management, monitoring and review of these patients, with clear benefits to emergency department staff, and potential benefits to patients and resource use.

**How this study might affect research, practice or policy:** SHarED illustrates the considerable potential for a quality improvement programme to promote more integrated case management by specialist teams across the health and care system for particularly vulnerable individuals and improve working arrangements for hard-pressed staff.

## INTRODUCTION

Frequent attenders or high-impact users (HIUs) are usually defined as patients who attend a hospital emergency department (ED) five or more times within 12 months [1]. These patients often have complex medical needs, are frequent users of other health and social care services including hospital admissions, and have higher mortality rates [1,2, 4,5,6]. In England, 2.1% of ED attenders presented five or more times in 2016/17, accounting for 10.7% of ED attendances [3]. The particular needs of HIUs, in addition to system-level challenges regarding management of urgent and emergency care pathways, including delays in hospital discharge, have contributed to EDs being a difficult work environment for staff [4,5].

In 2017, the Royal College of Emergency Medicine [1] and NHS England promoted action to ensure that HIUs have their needs “met more effectively through an improved, integrated service, reducing their future attendances at A&E” [1]. In 2022, NHS England announced that it was working to increase the provision of high-intensity user services [6,7]. The use of personalised care plans has been advocated for these patients [1,7,8], potentially reducing ED attendance and hospital admissions [4,9–12]. Despite national recognition of the rationale for better management of HIUs, service improvement has relied on the local efforts of clinicians to change working practices [10,13].

In 2020, the West of England Academic Health Science Network (AHSN) launched the Supporting High impAct useRs in Emergency Departments (SHarED) quality improvement programme to spread a HIU intervention first developed in the Bristol Royal Infirmary (BRI) ED across the other six EDs in five local hospital Trusts [10]. SHarED provided funding for ED staff to implement processes for the identification, proactive management, monitoring and review of HIUs [14].

To evaluate SHarED, we aimed to understand a range of factors including implementation, changes in working practices, impact, sustainability, and resource use, from the perspective of ED staff with HIU roles in the five participating Trusts. We sought to provide further insight on the impact of SHarED by analysing data collected during the programme. The findings are intended to inform the potential future rollout of an HIU quality improvement programme.

## METHODS

### Setting

All six EDs (in addition to the BRI’s ED) in the five hospital Trusts within the West of England AHSN participated: Gloucestershire Hospitals NHS Foundation Trust, Great Western Hospitals NHS Foundation Trust, North Bristol NHS Trust, Royal United Hospitals Bath NHS Foundation Trust, and University Hospitals Bristol and Weston NHS Foundation Trust.

### Intervention development and description

The HIU team at the BRI was formed in 2015 to “share specialised clinical knowledge to support individual patients to make better decisions about their health by addressing their specific issues and formulating a personal support plan” [10]. The initial focus was frequent attenders presenting with drug/alcohol problems and mental health issues, who were homeless, and whose behaviour, including verbal aggression, impacted staff [10]. Over its first 12 months, the BRI HIU team instigated multidisciplinary collaboration to create personal support plans (PSPs) for 87 selected HIUs, who subsequently experienced reductions in ED attendance and hospital admissions [10]. In 2017, the Trust funded a part-time HIU nurse co-ordinator and some of the lead ED consultant’s time to facilitate their HIU work.

SHarED broadly followed the collaborative quality improvement programme model developed by Berwick to promote the BRI’s HIU approach across the region [15]. SHarED aimed to reduce ED attendance of enrolled HIUs by 20% over 12 months and improve the experience of HIUs and ED staff over the period. In addition to the provision of clinical leadership, project management, and opportunities for peer-support and networking, SHarED created a toolkit to support each component of the intervention: i) referral, triaging and prioritisation of HIUs, ii) creation of PSPs, iii) arrangements for multi-disciplinary team meetings, iv) communication with patients and GPs, v) ED staff training and vi) data collection and reporting arrangements.

SHarED funded part-time roles for a HIU nurse coordinator and lead consultant in each participating ED, initially for 20 weeks, with an expectation that approximately 20 HIUs would be recruited during this period. The project was officially launched in 2019 but was delayed to 2020 due to the Covid-19 pandemic. Funding was also extended, in part due to constraints associated with the Covid-19 pandemic.

### Intervention evaluation

Our evaluation had two components. First, a qualitative study of views of participating HIU staff on the implementation of SHarED and its subsequent sustainability, which was informed by the realist evaluation framework to understand: how HIUs staff engaged with and implemented the SHarED, how working practices relating to HIUs changed during the funded period, the perceived impact of the SHarED programme, the barriers and facilitators relating to its sustainability, and how the SHarED funding was used to deliver programme [16]. Second, an analysis of anonymised data collected by the SHarED programme for service evaluation of: the number of ED attendances and hospital admissions for HIUs included in the SHarED programme, and estimated impact of changes on resource use; the number of ED staff trained in HIU management, and) responses from ED staff to three surveys on their experiences in managing HIUs.

### Data sources and measures

The SHarED project manager invited six HIU consultant leads, including the SHarED and BRI clinical lead, and five HIU coordinators to be interviewed. One consultant and one coordinator had taken on their HIU role after the SHarED had been completed. One of the HIU coordinators did not respond to the invitation, so the researcher undertook 10 interviews with consultant leads from all five participating Trusts and coordinators from four of the five Trusts. The interviews were undertaken between October and February 2022, lasting from 45-60 minutes and were conducted via Microsoft Teams. The interviews were guided by an iteratively developed topic guide, which covered themes including, history, planning and implementation, delivery, impact, sustainability, and resource use.

Anonymised quantitative data were collected by the project manager as part of the SHarED programme for service evaluation. Data for HIUs from the six participating EDs included age, Trust, and number of ED attendances and hospital admissions per month for 12 months before and after the intervention. The number of staff trained in HIU management during SHarED was also collected from each ED. The SHarED’s staff experience survey included 12 questions about managing HIUs and was sent to ED staff three times: September 2020, March 2021, and July 2021 (supplementary material Table S1).

For ED attendance we used the national average cost from the 2021/22 National Cost Collection for the NHS [17]. To estimate an average cost for a hospital admission applicable to HIUs we used internal data from the BRI for activity from Healthcare Resource Group codes applicable to this type of patient cohort, cost for each code was also retrieved from the National Cost Collection (see supplementary material Table S2).

### Analysis

Interview audio files were transcribed, anonymised and checked for accuracy. Data analysis was based on Framework methods, which apply a thematic structure to summarise and classify data using a series of matrices to organise data informed by the research questions, interview topic guide, and realist approach [18,19]. The text was coded to represent key topics/themes and recorded in columns, with rows representing individual informants. This facilitated the exploration of themes and patterns across the range of informants. Analysis of the first interviews informed further data collection: early insights informed changes to the topic guide used during later interviews. Researchers coded the interviews they conducted, independently double-coded a sub-sample of transcripts, and discussed the preliminary coding framework and themes to ensure the rigour of the emerging analysis.

The number of monthly ED attendances and hospital admissions for HIUs during the 12 months before and after being enrolled in SHarED were summarised using descriptive statistics. Due to the variation in the duration of follow-up data, we categorised HIUs by the number of months with follow-up data, from three to 12 months. Researchers analysed emergency visits and hospital admissions for each HIUs category using two baseline periods: i) 12 months before the intervention, ii) the number of months before enrolment to match the number of months of follow-up data. Differences in the average of monthly ED visits and hospital admissions were compared using t-tests for HIUs with follow-up periods of three, six, nine, and 12 months. The number and proportion of staff trained in EDs was reported. Staff survey data were summarised by the mean score, on a scale of 0-100, per question, by staff role and survey wave. Differences in mean scores between staff roles and waves were compared using t-tests. The open text questions were coded, each to different themes, reporting the frequencies that staff mentioned each theme for each wave. Statistical analysis was conducted in R 4.2.1 and Stata 16.

We report the SHarED funding and impact on ED attendance and hospital admission costs, based on the six month follow-up data, from a Trust perspective using local cost data.

### Ethical Considerations

The qualitative study was approved by the University of Bristol Faculty of Health Sciences Research Ethics Committee (approval code: 10275) and the NHS Health Research Authority and Health and Care Research Wales (IRAS project ID: 300027) and local R&D from the five Trusts. The analysis of the anonymised quantitative data collected as part of the SHarED programme for service evaluation was managed using a Data Protection Impact Assessment with the five Trusts.

## RESULTS

### HIU staff views

#### Working environment in EDs

The EDs were unanimously viewed as difficult and challenging environments for staff due to longstanding issues, including system-level pressures which had a negative impact on staff:

*Every year I think it can’t get any worse and every year it does. […] it’s just exceptionally busy, there’s lots of people who can’t access any other resources, so ED is where everyone comes at the end of the day, whether they need to or not so it’s busier and busier. (#8)*

#### HIU characteristics and challenges for EDs and the wider health and care system

HIUs were broadly characterised by high levels of vulnerability and marginalisation, facing a wide range of issues from mental health, alcohol and drug abuse, violence, exploitation, homelessness, and chaotic life conditions to chronic pain and complex medical conditions. The circumstances of HIUs when presenting at ED spanned a wide spectrum, including individuals widely viewed as being difficult for staff to manage in the ED. For these individuals, the standard protocols for assessment and treatment were not viewed as fit-for-purpose, resulting in challenging encounters for staff in the ED and more widely across the health and care system.

While SHarED’s initial HIU definition was those attending ED five or more times in the previous 12 months, the participating EDs typically implemented their criteria for identifying HIUs, including a focus on attendance over a shorter period. All the Trusts further broadened their approach to allow staff to refer individuals viewed as having a high impact on staff during an attendance:

*any member of staff, like reception, nurses, doctors, other people outside ED can refer […] it gives the staff something they feel they’re doing for those patients. So we find it quite advantageous because it makes people feel empowered to make a difference. (#7)*

#### The SHarED programme’s core characteristics and local implementation

SHarED enabled ED staff to learn about BRI’s HIU model, which provided a practical framework for improving the management of HIUs and could be adapted to local characteristics. The project management and materials provided, such as template letters to patients and other professionals and data recording tools, were viewed as useful. The AHSN funding enabled HIU nurse co-ordinator roles and consultant leads to be recruited with time dedicated to the project and the HIU Team. The networking opportunities with the other HIUs teams for staff participating in SHarED, during and after the programme, were highly valued. SHarED enabled previously unstructured and improvised HIU initiatives to evolve into more established and clearly defined services.

#### Proactive management of HIUs: staff roles in the ED and wider health and care system

Initial contact with the HIU or their GP by letter could lead to behaviour change clearly benefiting both the individual and the health care system. For more complex cases, the core element of HIU management was the creation, sharing and use of a personal support plan (PSP). Depending on the HIU’s circumstances, the PSP would be written and signed off by the HIU nurse co-ordinator, HIU consultant lead, or specialists from outside the ED. PSPs had three key objectives: i) engagement with HIUs about their behaviour and needs; ii) recommendations for ED clinicians to aid their interaction with HIUs, and iii) collaboration between professionals working across the health and care system to share specialist expertise and co-ordinate person-centred care:

*It’s a really important way for us to convey [*…*] really vital information to our clinicians at any time of day*. (#10)

Many PSPs had a key multidisciplinary component, such that specialist teams (mental health, safeguarding, pain, community, homelessness, drug and alcohol) either verified recommendations made by ED clinicians, led on generation of new recommendations, or shared their existing plans. One Trust’s HIU team included a pain consultant and a HIU co-ordinator based in its safeguarding team. This broader perspective on HIU staff roles represented a more holistic approach to managing HIUs, which actively promoted more integrated case management for HIUs across health and social care providers.

#### Benefits for ED patients, staff, and the wider health and care system

The HIU staff were confident HIUs benefited, although being such a heterogeneous group this was manifested in different ways, from support to change behaviour (e.g., attending ED less frequently), or improved access to other specialist services, to a more consistent, person-centred care, with potentially less waiting and fewer unnecessary investigations, delivered with compassion:

*I expect that they feel a bit less stigma, so they’re not being left for longer than other patients in the waiting room, they’re not being treated any differently*. (#5)

In the context of the high turnover of medical and nursing staff, the use of PSPs was unanimously viewed as beneficial for frontline staff who felt more empowered when providing care to HIUs. The practical aid PSPs provided staff, and the raised awareness of HIUs’ circumstances, contributed to positive changes in culture relating to staff perceptions of HIUs, and tangibly reduced workload-related pressure. The more structured approach to an ED-based HIU service, facilitated by SHarED, enabled improved communication between different care providers and services to develop more holistic and integrated care across the health and care system.

*[Having a PSP] makes people feel more satisfied with an interaction and also more confident to go and see somebody for whom they don’t have the answer because they then have […] a plan where they know that these things have already been done*. (#4)

#### Impact on resource use

All the interviewed ED staff were confident that their HIU service had led to better care provision associated with a reduction in hospital resources used. However, the focus on quantifying impact in terms of changes in the number of ED attendances and hospital admissions was viewed as providing, in isolation, only limited insight into the impact of the HIU service. Although each HIU service generated a business case for funding to continue the SHarED intervention, only one ED was successful in securing long term funding through its Trust. One of the key issues for the business cases was a requirement to demonstrate cost savings. However, there appeared to be no consensus about what evidence would be viewed as sufficient to secure the level of funding required to continue the staff capacity enabled by the SHarED programme. One ED secured funding from its Integrated Care Board (ICB), rather than Trust, building on the existing wider HIU infrastructure already in place. Another ED secured funding from the existing ED budget.

#### Barriers and challenges

SHarED and HIUs teams faced challenges including recruiting and promoting roles for two HIUs teams, aligning goals with other programmes that could also support HIUs, and engaging clinical leads, management and ICBs in spite of being essential for HIUs teams performance and its sustainability. The supporting network between HIUs teams supported by SHarED was reported to be valuable; HIUs staff subsequently struggled to maintain this network without the AHSN support. All EDs experienced difficulties with securing funding to sustain their HIU service.

*We would all like to think that things that improve patient experience and staff experience should be valued and resourced by the NHS, but of course it often just comes down to finances. So, it would be rare that an idea just about staff welfare, or just about patient experience, would be funded*. (#5)

### Impact on ED attendance and hospital admissions

Three month post-enrolment follow-up data were reported for 148 HIUs from the six EDs. The availability of follow-up data reduced over time such that six month follow-up data were available for 86% (127/148) of cases, and for 22% (33/148) of cases at 12 months (Tables 1 and 2). During the 12 months prior to enrolment in SHarED, the mean number of ED attendances and hospital admissions per HIU increased over time (Figure 1). This pattern of activity reversed post-enrolment, with the reduction experienced during the first three months subsequently maintained, at least for those HIUs for whom further follow-up data were available (Figure 1).

**Table 1.** Mean monthly attendances per HIU enrolled in SHarED across the six participating EDs

| Months of follow-up | Number of HIUs (n) | Mean monthly number of ED visits pre- and post-enrolment in SHarED |  |  |  |  |  | Difference in the monthly number of ED visits |  |  |  |  |  |  |  |
| --- | --- | --- | --- | --- | --- | --- | --- | --- | --- | --- | --- | --- | --- | --- | --- |
|  |  | baseline 12 months |  | baseline equal to follow-up period |  | Follow-up |  | 12 months baseline |  |  |  | Baseline equal to follow-up period |  |  |  |
|  |  | Mean | SD | Mean | SD | Mean | SD | Value | 95%CI | p-value | Change (%) | Value | 95%CI | p-value | Change (%) |
| 3 | 148 | 1.6 | 1.0 | 2.4 | 2.1 | 1.6 | 1.9 | 0.0 | -0.29 to 0.30 | 0.991 | 0 | -0.8 | -1.12 to -0.41 | <0.001 | -33 |
| 4 | 142 | 1.6 | 1.0 | 2.3 | 1.7 | 1.5 | 1.8 |  |  |  |  |  |  |  |  |
| 5 | 135 | 1.7 | 1.0 | 2.2 | 1.5 | 1.5 | 1.6 |  |  |  |  |  |  |  |  |
| 6 | 127 | 1.6 | 1.0 | 2.1 | 1.3 | 1.4 | 1.5 | -0.3 | -0.55 to -0.03 | 0.030 | -13 | -0.7 | -0.98 to -0.43 | <0.001 | -33 |
| 7 | 116 | 1.7 | 1.0 | 2.0 | 1.2 | 1.3 | 1.4 |  |  |  |  |  |  |  |  |
| 8 | 110 | 1.7 | 1.0 | 1.9 | 1.1 | 1.3 | 1.4 |  |  |  |  |  |  |  |  |
| 9 | 96 | 1.8 | 1.0 | 1.9 | 1.0 | 1.3 | 1.4 | -0.5 | -0.72 to -0.17 | 0.002 | -28 | -0.6 | -0.88 to -0.34 | <0.001 | -32 |
| 10 | 81 | 1.8 | 1.0 | 1.9 | 1.1 | 1.3 | 1.4 |  |  |  |  |  |  |  |  |
| 11 | 45 | 1.5 | 0.9 | 1.5 | 0.9 | 1.1 | 1.2 |  |  |  |  |  |  |  |  |
| 12 | 33 | 1.6 | 1.0 | 1.6 | 1.0 | 1.0 | 1.3 | -0.5 | -0.93 to -0.13 | 0.011 | -38 | -0.5 | -0.93 to -0.13 | 0.011 | -38 |

**Table 2.** Mean monthly hospital admissions per HIU enrolled in SHarED across the six participating EDs

| Months of follow-up | Number of HIUs (n) | Mean monthly number of ED visits pre- and post-enrolment in SHarED |  |  |  |  |  | Difference in the monthly number of ED visits |  |  |  |  |  |  |  |
| --- | --- | --- | --- | --- | --- | --- | --- | --- | --- | --- | --- | --- | --- | --- | --- |
|  |  | 12 months baseline |  | Baseline equivalent to follow-up period |  | Follow-up |  | 12 months baseline |  |  |  | Baseline equivalent to follow-up period |  |  |  |
|  |  | Mean | SD | Mean | SD | Mean | SD | Value | 95%CI | p-value | Change (%) | Value | 95%CI | p-value | Change (%) |
| 3 | 148 | 0.5 | 0.5 | 0.6 | 0.8 | 0.3 | 0.5 | -0.2 | -0.25 to -0.09 | <0.001 | -40 | -0.3 | -0.44 to -0.19 | <0.001 | -50 |
| 4 | 142 | 0.5 | 0.5 | 0.6 | 0.7 | 0.3 | 0.4 |  |  |  |  |  |  |  |  |
| 5 | 135 | 0.5 | 0.5 | 0.6 | 0.6 | 0.3 | 0.4 |  |  |  |  |  |  |  |  |
| 6 | 127 | 0.5 | 0.5 | 0.6 | 0.6 | 0.2 | 0.3 | -0.3 | -0.33 to -0.17 | <0.001 | -60 | -0.3 | -0.41 to -0.24 | <0.001 | -67 |
| 7 | 116 | 0.5 | 0.5 | 0.6 | 0.6 | 0.2 | 0.3 |  |  |  |  |  |  |  |  |
| 8 | 110 | 0.5 | 0.5 | 0.6 | 0.6 | 0.2 | 0.3 |  |  |  |  |  |  |  |  |
| 9 | 96 | 0.6 | 0.5 | 0.6 | 0.6 | 0.2 | 0.3 | -0.4 | -0.44 to -0.25 | <0.001 | -67 | -0.4 | -0.47 to -0.27 | <0.001 | -67 |
| 10 | 81 | 0.6 | 0.5 | 0.6 | 0.6 | 0.2 | 0.3 |  |  |  |  |  |  |  |  |
| 11 | 45 | 0.4 | 0.4 | 0.4 | 0.4 | 0.2 | 0.3 |  |  |  |  |  |  |  |  |
| 12 | 33 | 0.4 | 0.4 | 0.4 | 0.4 | 0.2 | 0.2 | -0.2 | -0.37 to -0.09 | 0.002 | -50 | -0.2 | -0.39 to -0.09 | 0.002 | -50 |

**Figure 1.**
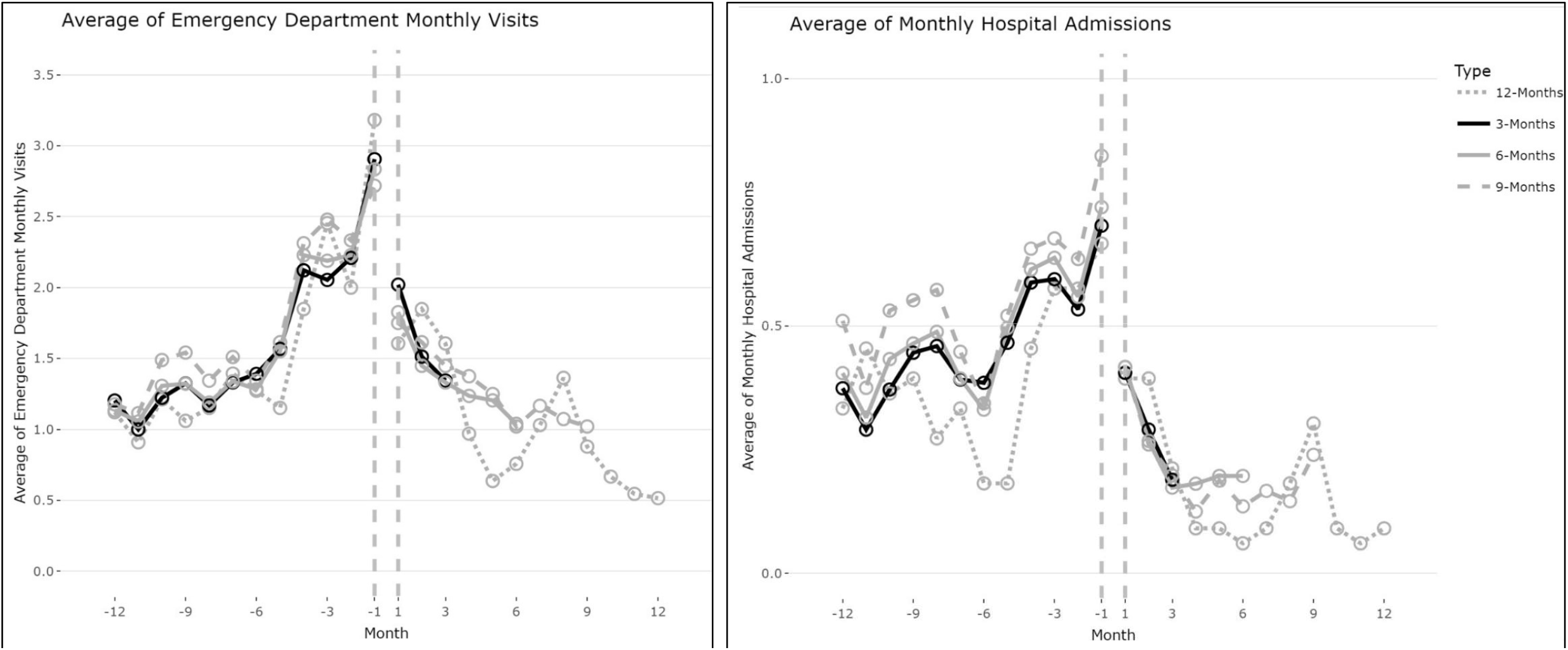
Mean monthly ED attendances and hospital admissions per HIU enrolled in SHarED across the six participating EDs Average of Emergency Department monthly visits (on the left) and the average of monthly hospital admissions (on the right) twelve months before personal supports plans were created and twelve months after for high-impact users categorised in four groups regarding the numbers of months with follow-up data: three, six, nine or twelve months.

Given the availability of follow-up data, comparison of the experience six months pre- and post-enrolment offers the best measure of the impact of SHarED: the mean monthly ED attendances per HIU reduced by 33% from 2.1 to 1.4, which was statistically significant (p<0.001) (Table 1) with a range from 27% to 49% between EDs. Between the same six-month periods, the mean monthly hospital admissions per HIU reduced by 67% from 0.6 to 0.2, which was statistically significant (p<0.001) (Table 2) with a range from 41% to 72% between EDs. A similar pattern of change over time was evident at three, nine and 12 months (Tables 1 and 2). The mean age of the 148 HIUs was 39.3 years (SD 15.7).

### ED staff training

During SHarED, 55% (372/671) of staff across the six participating EDs received training on the management of HIUs.

### ED staff survey

The survey of ED staff had response rates of 24% (258/1042) in wave one, 36% (348/967) in wave two and 12% (131/1092) in wave three. The survey included seven statements for which staff were asked to give a rating on a scale of 0 ‘strongly disagree’ to 100 ‘strongly agree’ (see Table S1 for statement text). Wave 1 represents baseline views and the subsequent waves indicate how views changed over time.

On average, staff who responded to the survey reported being more confident when assessing and treating HIUs over time: there was a clear evidence of the increase from 65 to 72 between waves one and three (p=0.002) (Table 3). Staff reported being similarly better supported when managing HIUs (Table 3). The largest improvement in ratings related to the statement ‘The department does a good job in training staff in how to manage High Impact Users’, which increased from 40 in wave one to 62 in both waves two and three (Table 3). The response to the statement ‘High Impact Users impact negatively on my mental wellbeing, either in the present or past’ did not significantly change over time. Statements relating to the care of HIUs, in terms of being treated with dignity and care, receiving appropriate assessment and treatment, and timely care, all received higher mean ratings over time (Table 3).

**Table 3.**
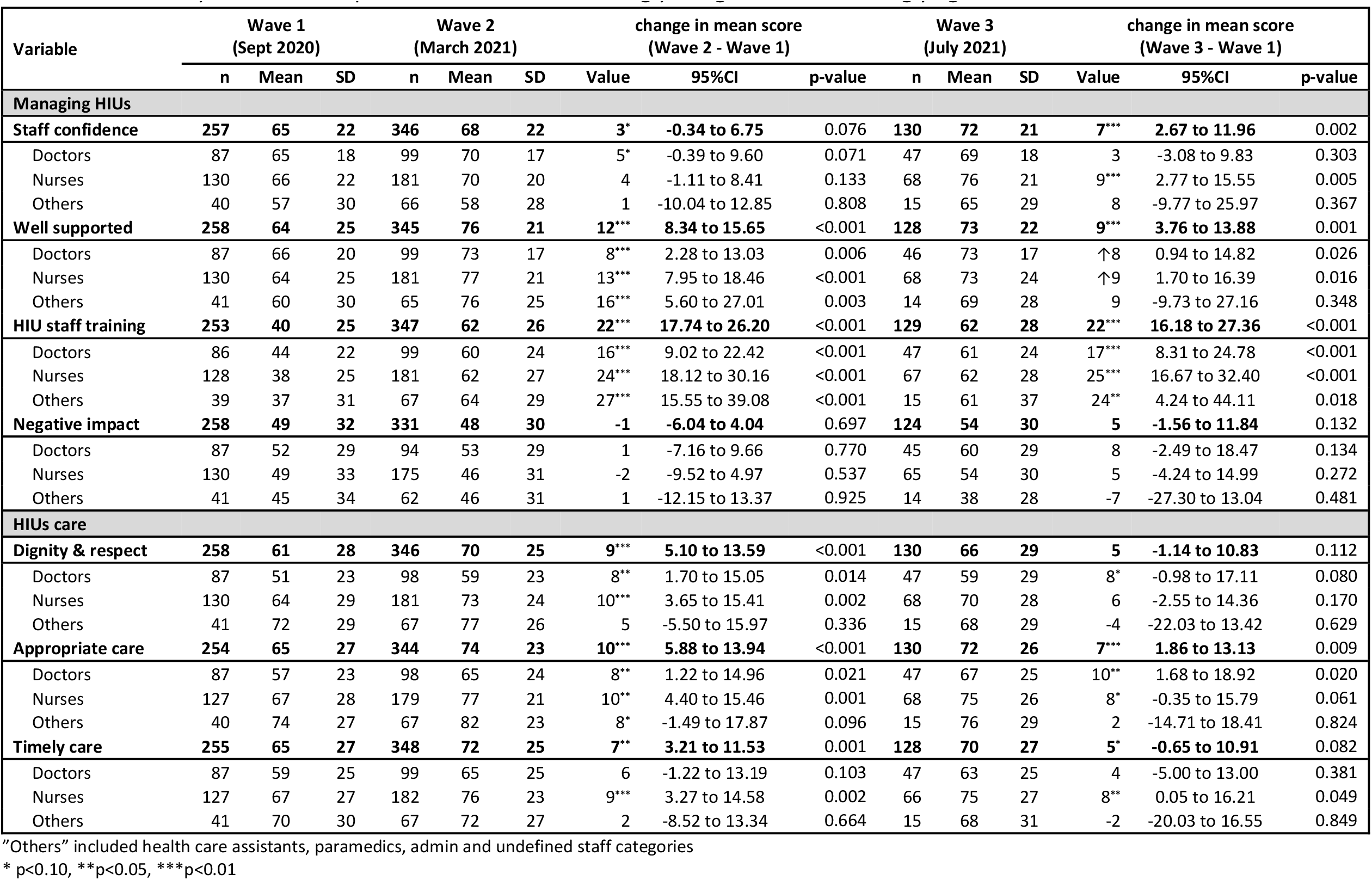
Staff survey statement responses on a scale of 0 ‘strongly disagree’ to 100 ‘strongly agree’

| Variable | Wave 1<br>(Sept 2020) |  |  | Wave 2<br>(March 2021) |  |  | change in mean score<br>(Wave 2 - Wave 1) |  |  | Wave 3<br>(July 2021) |  |  | change in mean score<br>(Wave 3 - Wave 1) |  |  |
| --- | --- | --- | --- | --- | --- | --- | --- | --- | --- | --- | --- | --- | --- | --- | --- |
|  | n | Mean | SD | n | Mean | SD | Value | 95%CI | p-value | n | Mean | SD | Value | 95%CI | p-value |
| **Managing HIUs** |  |  |  |  |  |  |  |  |  |  |  |  |  |  |  |
| **Staff confidence** | **257** | **65** | **22** | **346** | **68** | **22** | **3\*** | **-0.34 to 6.75** | 0.076 | **130** | **72** | **21** | **7\*\*\*** | **2.67 to 11.96** | 0.002 |
| Doctors | 87 | 65 | 18 | 99 | 70 | 17 | 5* | -0.39 to 9.60 | 0.071 | 47 | 69 | 18 | 3 | -3.08 to 9.83 | 0.303 |
| Nurses | 130 | 66 | 22 | 181 | 70 | 20 | 4 | -1.11 to 8.41 | 0.133 | 68 | 76 | 21 | 9*** | 2.77 to 15.55 | 0.005 |
| Others | 40 | 57 | 30 | 66 | 58 | 28 | 1 | -10.04 to 12.85 | 0.808 | 15 | 65 | 29 | 8 | -9.77 to 25.97 | 0.367 |
| **Well supported** | **258** | **64** | **25** | **345** | **76** | **21** | **12\*\*\*** | **8.34 to 15.65** | <0.001 | **128** | **73** | **22** | **9\*\*\*** | **3.76 to 13.88** | 0.001 |
| Doctors | 87 | 66 | 20 | 99 | 73 | 17 | 8*** | 2.28 to 13.03 | 0.006 | 46 | 73 | 17 | ↑8 | 0.94 to 14.82 | 0.026 |
| Nurses | 130 | 64 | 25 | 181 | 77 | 21 | 13*** | 7.95 to 18.46 | <0.001 | 68 | 73 | 24 | ↑9 | 1.70 to 16.39 | 0.016 |
| Others | 41 | 60 | 30 | 65 | 76 | 25 | 16*** | 5.60 to 27.01 | 0.003 | 14 | 69 | 28 | 9 | -9.73 to 27.16 | 0.348 |
| **HIU staff training** | **253** | **40** | **25** | **347** | **62** | **26** | **22\*\*\*** | **17.74 to 26.20** | <0.001 | **129** | **62** | **28** | **22\*\*\*** | **16.18 to 27.36** | <0.001 |
| Doctors | 86 | 44 | 22 | 99 | 60 | 24 | 16*** | 9.02 to 22.42 | <0.001 | 47 | 61 | 24 | 17*** | 8.31 to 24.78 | <0.001 |
| Nurses | 128 | 38 | 25 | 181 | 62 | 27 | 24*** | 18.12 to 30.16 | <0.001 | 67 | 62 | 28 | 25*** | 16.67 to 32.40 | <0.001 |
| Others | 39 | 37 | 31 | 67 | 64 | 29 | 27*** | 15.55 to 39.08 | <0.001 | 15 | 61 | 37 | 24** | 4.24 to 44.11 | 0.018 |
| **Negative impact** | **258** | **49** | **32** | **331** | **48** | **30** | **-1** | **-6.04 to 4.04** | 0.697 | **124** | **54** | **30** | **5** | **-1.56 to 11.84** | 0.132 |
| Doctors | 87 | 52 | 29 | 94 | 53 | 29 | 1 | -7.16 to 9.66 | 0.770 | 45 | 60 | 29 | 8 | -2.49 to 18.47 | 0.134 |
| Nurses | 130 | 49 | 33 | 175 | 46 | 31 | -2 | -9.52 to 4.97 | 0.537 | 65 | 54 | 30 | 5 | -4.24 to 14.99 | 0.272 |
| Others | 41 | 45 | 34 | 62 | 46 | 31 | 1 | -12.15 to 13.37 | 0.925 | 14 | 38 | 28 | -7 | -27.30 to 13.04 | 0.481 |
| **HIUs care** |  |  |  |  |  |  |  |  |  |  |  |  |  |  |  |
| **Dignity & respect** | **258** | **61** | **28** | **346** | **70** | **25** | **9\*\*\*** | **5.10 to 13.59** | <0.001 | **130** | **66** | **29** | **5** | **-1.14 to 10.83** | 0.112 |
| Doctors | 87 | 51 | 23 | 98 | 59 | 23 | 8** | 1.70 to 15.05 | 0.014 | 47 | 59 | 29 | 8* | -0.98 to 17.11 | 0.080 |
| Nurses | 130 | 64 | 29 | 181 | 73 | 24 | 10*** | 3.65 to 15.41 | 0.002 | 68 | 70 | 28 | 6 | -2.55 to 14.36 | 0.170 |
| Others | 41 | 72 | 29 | 67 | 77 | 26 | 5 | -5.50 to 15.97 | 0.336 | 15 | 68 | 29 | -4 | -22.03 to 13.42 | 0.629 |
| **Appropriate care** | **254** | **65** | **27** | **344** | **74** | **23** | **10\*\*\*** | **5.88 to 13.94** | <0.001 | **130** | **72** | **26** | **7\*\*\*** | **1.86 to 13.13** | 0.009 |
| Doctors | 87 | 57 | 23 | 98 | 65 | 24 | 8** | 1.22 to 14.96 | 0.021 | 47 | 67 | 25 | 10** | 1.68 to 18.92 | 0.020 |
| Nurses | 127 | 67 | 28 | 179 | 77 | 21 | 10** | 4.40 to 15.46 | 0.001 | 68 | 75 | 26 | 8* | -0.35 to 15.79 | 0.061 |
| Others | 40 | 74 | 27 | 67 | 82 | 23 | 8* | -1.49 to 17.87 | 0.096 | 15 | 76 | 29 | 2 | -14.71 to 18.41 | 0.824 |
| **Timely care** | **255** | **65** | **27** | **348** | **72** | **25** | **7\*\*** | **3.21 to 11.53** | 0.001 | **128** | **70** | **27** | **5\*** | **-0.65 to 10.91** | 0.082 |
| Doctors | 87 | 59 | 25 | 99 | 65 | 25 | 6 | -1.22 to 13.19 | 0.103 | 47 | 63 | 25 | 4 | -5.00 to 13.00 | 0.381 |
| Nurses | 127 | 67 | 27 | 182 | 76 | 23 | 9*** | 3.27 to 14.58 | 0.002 | 66 | 75 | 27 | 8** | 0.05 to 16.21 | 0.049 |
| Others | 41 | 70 | 30 | 67 | 72 | 27 | 2 | -8.52 to 13.34 | 0.664 | 15 | 68 | 31 | -2 | -20.03 to 16.55 | 0.849 |

"Others" included health care assistants, paramedics, admin and undefined staff categories
\* p<0.10, \*\*p<0.05, \*\*\*p<0.01

We assigned responses to the survey’s open-text questions to themes, which are summarised in Figure 2. ED staff were more aware of how to refer patients to their HIU service over time and that the PSPs were increasingly available via electronic patient records. Staff reported being more sure about the care provided to HIUs, but were also more aware of the risk of violence from these patients. Staff also reported that the major risks to HIUs were the provision of inappropriate assessment or management, and missed diagnoses (Figure 2).

**Figure 2.**
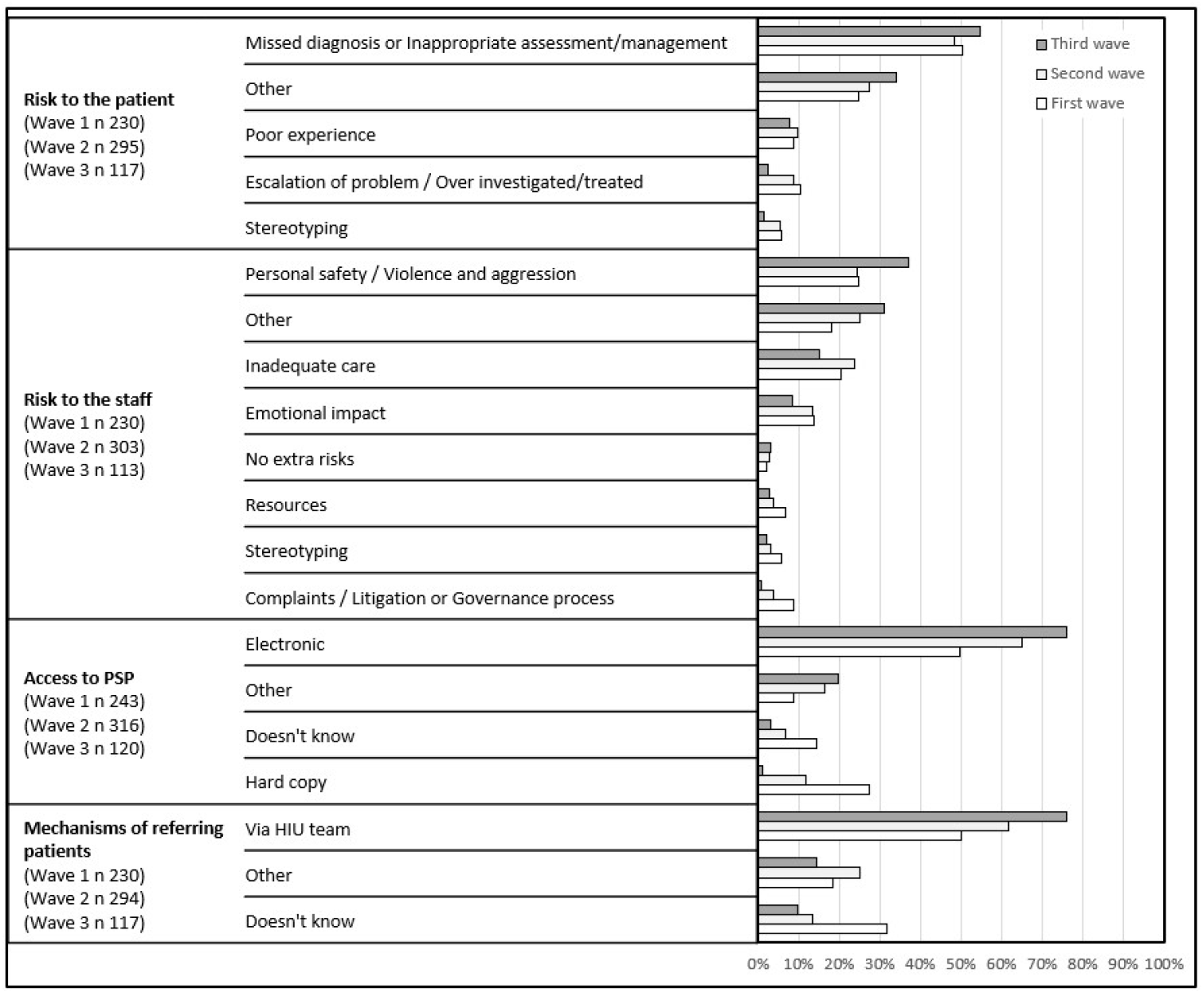
Staff survey themes assigned to open text question responses.

### Estimated impact on resources

SHarED costs totalled £139,944 including project management (£46,453), BRI time (£23,679) and funding to EDs for staff time (£69,812). Using the six-month follow-up experience, there was a reduction of 528 ED visits and 306 hospital admissions. The estimated mean costs of ED attendance and hospital admissions avoided were £297 and £577, respectively. Therefore, the estimated impact on costs associated with the reduction in ED attendance and hospital admissions was £333,529. Deducting the SHarED costs suggested a potential net cost saving of up to £193,585. If this change in activity is assumed to have also have been experienced by the 21 HIUs for whom six month follow-up data were not available, the total cost saving was £388,680 with a net cost saving of £248,736.

## DISCUSSION

SHarED responded to a nationally recognised need to improve the management of HIUs by delivering a quality improvement programme to promote one ED’s established model across the other six EDs in the region. SHarED was the first ED-based HIU quality improvement programme in the NHS, and it was successful in enabling clinicians to establish more person-centred services for HIUs, despite the concurrent challenges associated with Covid-19. SHarED aimed to reduce ED attendance by 20% over 12 months for selected HIUs and the available data indicate that this level of impact was achieved by all sites with an estimated consequent positive impact on resource use.

The ED clinicians with HIU roles were unanimously positive about the benefits of SHarED for both staff and patients, and the staff survey demonstrated a wider impact on issues including awareness and training. A key component of the HIU model was the use of personal support plans to strengthen both care planning and case management. SHarED proactive management of HIUs enabled better care planning by supporting frontline ED staff with patient-centred recommendations for HIUs and provided the basis for more integrated case management by specialist teams across the health and care system [4,8,12,20–22]. One of the participating Trusts contributed a key role in promoting this more holistic approach, which was enabled by leadership from the local ICB, and could be viewed as an exemplar for future development of a future quality improvement programme. Moreover, frequent ED attendance can be viewed as characteristic of a complex system [22], which requires solutions at a system level as well as at the individual level [22,23], thus, a system-wide approach has been recommended [13,24].

SHarED participants’ locally determined criteria for identifying HIUs mirrored the lack of a single definition of HIUs in the literature [25–27], as local differences in population characteristics and services influence the heterogeneity of this diverse group of individuals with complex health needs [2,3,12,24–26,28,29].

The SHarED programme, through the establishment of HIUs teams and, more specifically through the use of PSPs, improved the working conditions and workload of frontline ED staff who were more able to provide appropriate and compassionate care and better manage of HIUs’ expectations. Indeed, SHarED contributed to a cultural shift in ED staff toward this vulnerable group of patients that reported to have experienced stigma and discriminatory behaviour from healthcare staff before [13,30,31].

Our evaluation of the impact of SHarED on resource use is limited because data on ED attendance and hospital admissions were collected for only a small number of HIUs as staff implemented new ways of working. Some reduction in resource use by HIUs would be expected due to mortality, moving away, incarceration, or other unrelated resolution of needs. More robust evidence of the impact of this type of quality improvement programme on HIU resource use could be generated by analysing a range of routine data using more sophisticated designs. While one current study will contribute to this [32], the evidence presented here warrants consideration. If we assume SHarED may have been responsible for half the reduction in ED attendance and hospital admissions, it would have generated a net cost saving of approximately £54,396 for the 148 HIUs.

### Suggested next steps

Our evaluation provides a basis for developing the SHarED programme for more widespread use. SHarED focused on improving the delivery of person-centred care for HIUs and also improved the working conditions and workload of frontline ED staff, which is particularly important during this sustained period of national crisis in the delivery of emergency medicine.

Facilitation of ongoing support for the cross-organisational networking and peer-support opportunities for staff would enable further valuable learning to promote HIU service sustainability and innovation, for ICB commissioners, ED staff, Trust leaders and wider stakeholders.

Giving SHarED a more holistic orientation to reflect the wider impact of HIUs on the health and care system would be advantageous. One option for achieving would be to seek explicit engagement from ICB commissioners, in addition to ED staff and acute Trust leaders, when recruiting new localities.

Additional support for HIU staff to make the case for further funding from Trusts and/or ICBs would be warranted to sustain the new working practices. Despite the confidence of participating staff that their HIU services had led to better care provision associated with a reduction in the use of hospital resources, there was no consensus about what evidence would be viewed as sufficient to secure ongoing funding for the type of HIU service enabled by the SHarED programme.

## CONCLUSIONS

Overall, SHarED illustrates the considerable potential for a quality improvement programme to promote more integrated case management by specialist teams across the health and care system for high-impact users as particularly vulnerable individuals, and improve working arrangements for hard-pressed ED staff. Health and care system bodies should consider positively the funding and sustaining of high-impact user teams in emergency departments and the widespread of SHarED quality improvement.

## Data Availability

In order to preserve the anonymity of the interview participants, the interview data will not
be made available. The rest of the data related to this study will be available upon reasonable request.

## SUPPLEMENTARY MATERIAL

**Table S1**. Staff survey questions

**Table S2**. Bristol Royal Infirmary activity for Health Research Groups codes applicable to High-Impact Users, and National Cost Collection for the NHS 2021/2022

## Acknowledgements

The project team would like to thank Steve Strong and Sarah Biggs who provided helpful feedback on our qualitative analysis from a public perspective perspective.

## Contributors

CSR and HM as researchers conceptualised and designed the study and data collection method to study high-impact user staff views; conducted semi-structured interviews and carried out the analyses. MK as the project manager of SHarED contributed in recruiting participants for interviews. WEHANS designed the data collection method and collected data for HIU attendances, hospital admissions, number of staff trained and staff survey for the purpose of service evaluation; CSR and HM analysed this data. RT provided the clinical leadership for SHarED. CSR and HM drafted the initial manuscript, reviewed and revised the manuscript. All authors and reviewed and revised the manuscript. All authors approved the final manuscript as submitted and agree to be accountable for all aspects of the work.

## Data availability statement

In order to preserve the anonymity of the interview participants, the interview data will not be made available.

---

**Table S1.**
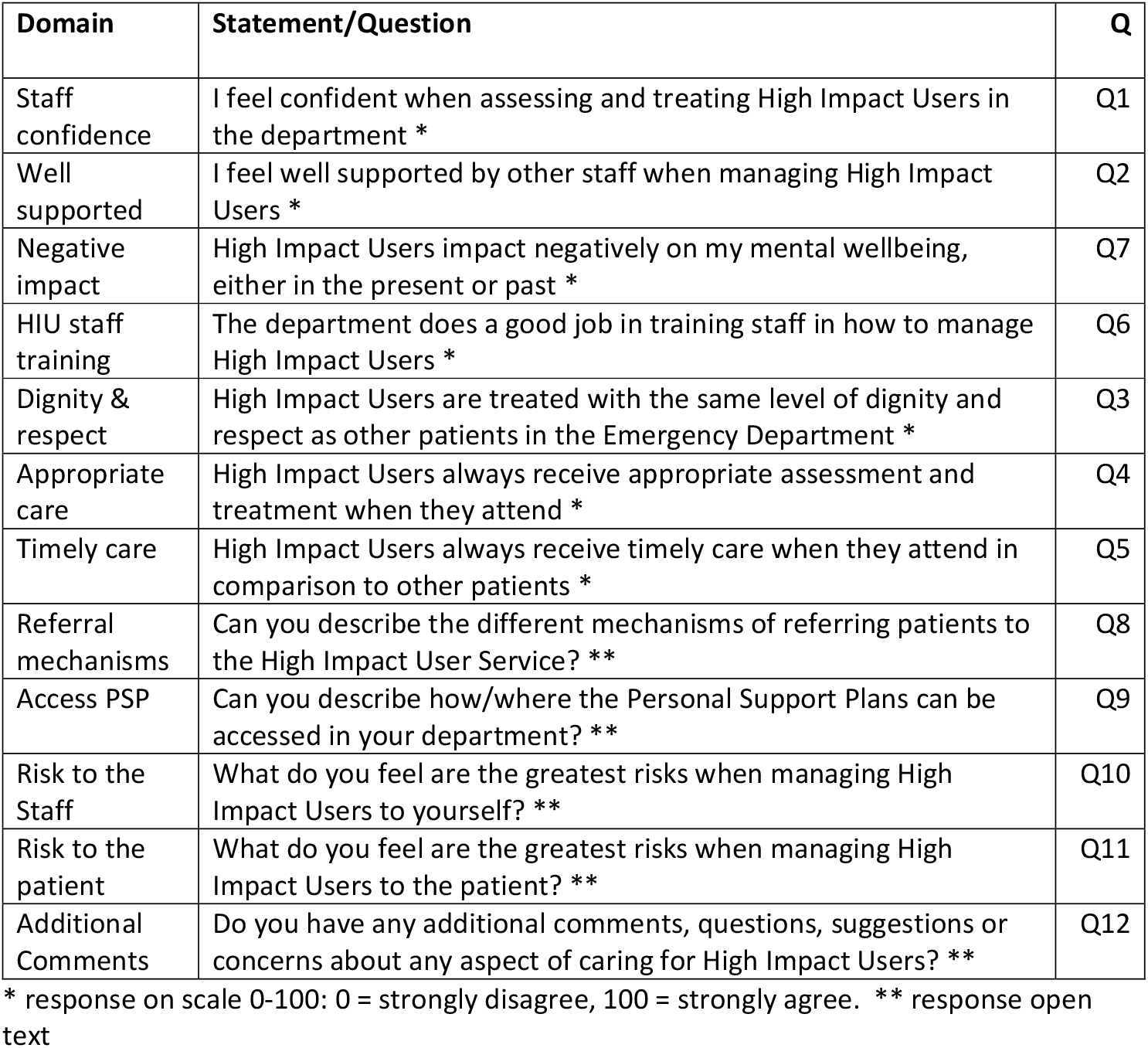
Staff survey questions

**Table S2.**
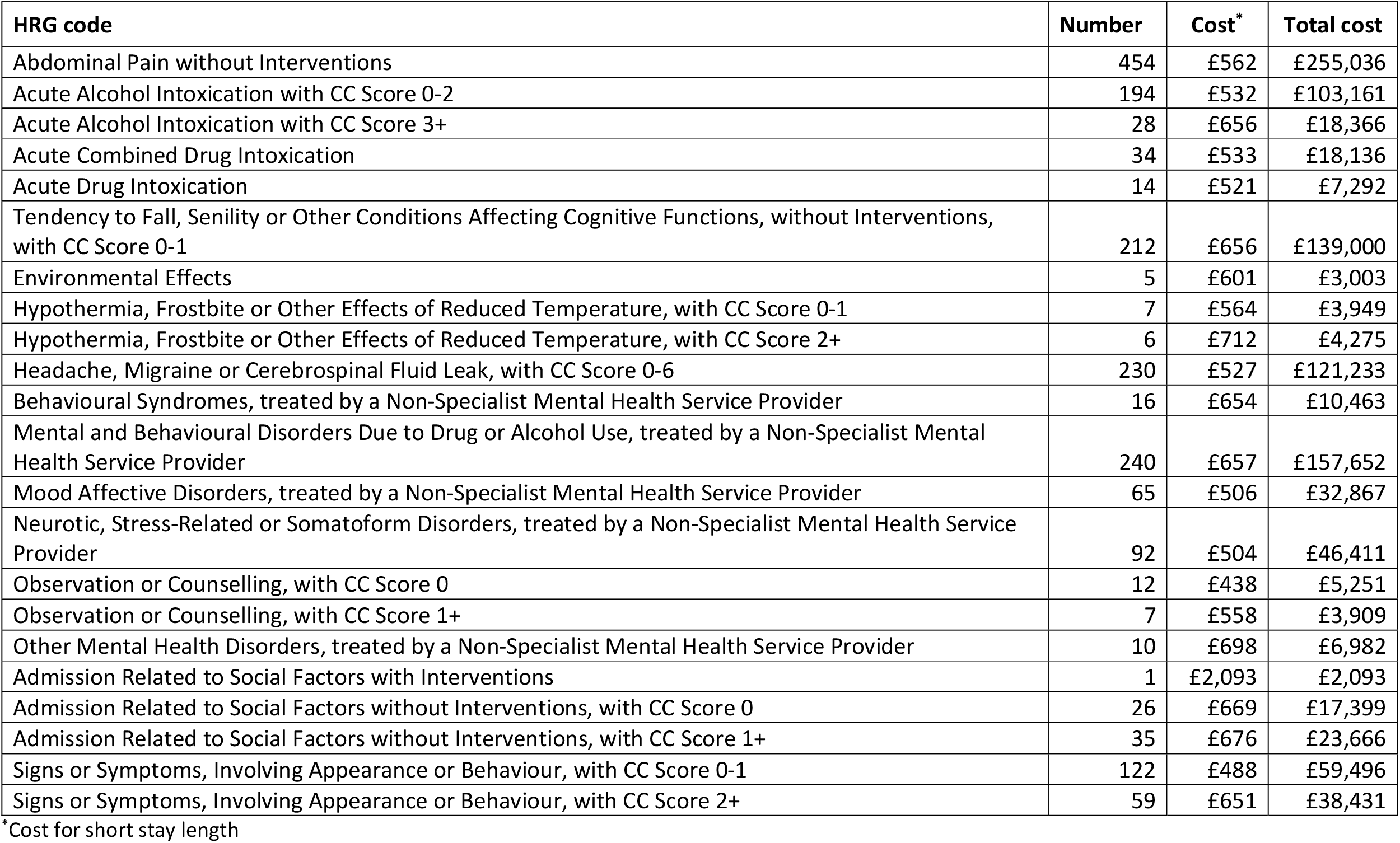
Bristol Royal Infirmary activity for Health Research Groups codes applicable to High-Impact Users, and National Cost Collection for the NHS 2021/2022

