## Supplementary Material for "Improving care for high impact users of hospital emergency departments: a mixed-method evaluation of a regional quality improvement programme ‘Supporting High impact users in the Emergency Department’ (SHarED)"

**Table S1**. Staff survey questions

| Domain | Statement/Question | Q |
|---|---|---|
| Staff confidence | I feel confident when assessing and treating High Impact Users in the department * | Q1 |
| Well supported | I feel well supported by other staff when managing High Impact Users * | Q2 |
| Negative impact | High Impact Users impact negatively on my mental wellbeing, either in the present or past * | Q7 |
| HIU staff training | The department does a good job in training staff in how to manage High Impact Users * | Q6 |
| Dignity & respect | High Impact Users are treated with the same level of dignity and respect as other patients in the Emergency Department * | Q3 |
| Appropriate care | High Impact Users always receive appropriate assessment and treatment when they attend * | Q4 |
| Timely care | High Impact Users always receive timely care when they attend in comparison to other patients * | Q5 |
| Referral mechanisms | Can you describe the different mechanisms of referring patients to the High Impact User Service? ** | Q8 |
| Access PSP | Can you describe how/where the Personal Support Plans can be accessed in your department? ** | Q9 |
| Risk to the Staff | What do you feel are the greatest risks when managing High Impact Users to yourself? ** | Q10 |
| Risk to the patient | What do you feel are the greatest risks when managing High Impact Users to the patient? ** | Q11 |
| Additional Comments | Do you have any additional comments, questions, suggestions or concerns about any aspect of caring for High Impact Users? ** | Q12 |

* response on scale 0-100: 0 = strongly disagree, 100 = strongly agree.  ** response open text

**Table S2**. Bristol Royal Infirmary activity for Health Research Groups codes applicable to High-Impact Users, and National Cost Collection for the NHS 2021/2022

| HRG code | Number | Cost* | Total cost |
|---|---|---|---|
| Abdominal Pain without Interventions | 454 | £562 | £255,036 |
| Acute Alcohol Intoxication with CC Score 0-2 | 194 | £532 | £103,161 |
| Acute Alcohol Intoxication with CC Score 3+ | 28 | £656 | £18,366 |
| Acute Combined Drug Intoxication | 34 | £533 | £18,136 |
| Acute Drug Intoxication | 14 | £521 | £7,292 |
| Tendency to Fall, Senility or Other Conditions Affecting Cognitive Functions, without Interventions, with CC Score 0-1 | 212 | £656 | £139,000 |
| Environmental Effects | 5 | £601 | £3,003 |
| Hypothermia, Frostbite or Other Effects of Reduced Temperature, with CC Score 0-1 | 7 | £564 | £3,949 |
| Hypothermia, Frostbite or Other Effects of Reduced Temperature, with CC Score 2+ | 6 | £712 | £4,275 |
| Headache, Migraine or Cerebrospinal Fluid Leak, with CC Score 0-6 | 230 | £527 | £121,233 |
| Behavioural Syndromes, treated by a Non-Specialist Mental Health Service Provider | 16 | £654 | £10,463 |
| Mental and Behavioural Disorders Due to Drug or Alcohol Use, treated by a Non-Specialist Mental Health Service Provider | 240 | £657 | £157,652 |
| Mood Affective Disorders, treated by a Non-Specialist Mental Health Service Provider | 65 | £506 | £32,867 |
| Neurotic, Stress-Related or Somatoform Disorders, treated by a Non-Specialist Mental Health Service Provider | 92 | £504 | £46,411 |
| Observation or Counselling, with CC Score 0 | 12 | £438 | £5,251 |
| Observation or Counselling, with CC Score 1+ | 7 | £558 | £3,909 |
| Other Mental Health Disorders, treated by a Non-Specialist Mental Health Service Provider | 10 | £698 | £6,982 |
| Admission Related to Social Factors with Interventions | 1 | £2,093 | £2,093 |
| Admission Related to Social Factors without Interventions, with CC Score 0 | 26 | £669 | £17,399 |
| Admission Related to Social Factors without Interventions, with CC Score 1+ | 35 | £676 | £23,666 |
| Signs or Symptoms, Involving Appearance or Behaviour, with CC Score 0-1 | 122 | £488 | £59,496 |
| Signs or Symptoms, Involving Appearance or Behaviour, with CC Score 2+ | 59 | £651 | £38,431 |

*Cost for short stay length
